# A 29-mRNA Host Response Test from Blood Accurately Distinguishes Bacterial and Viral Infections Among Emergency Department Patients

**DOI:** 10.1101/2020.12.01.20242321

**Authors:** Asimina Safarika, James W. Wacker, Konstantinos Katsaros, Nicky Solomonidi, George Giannikopoulos, Antigone Kostaki, Ioannis M. Koutelidakis, Sabrina M. Coyle, Henry K. Cheng, Oliver Liesenfeld, Timothy E. Sweeney, Evangelos J. Giamarellos-Bourboulis

**Affiliations:** 4th Department of Internal Medicine, National and Kapodistrian University of Athens, Greece; Inflammatix Inc, Clinical Affairs, Burlingame, CA, United States; Department of Surgery, Nafplion General Hospital, Greece; Department of Internal Medicine, Syros General Hospital, Greece; 2^nd^ Department of Surgery, Aristotle University of Thessaloniki, Greece

**Keywords:** Acute infection, sepsis, host response, diagnostics, gene expression, InSep, Emergency Department

## Abstract

**Study design:** Whether or not to administer antibiotics is a common and challenging clinical decision in patients with suspected infections presenting to the emergency department (ED). We prospectively validate InSep, a 29-mRNA blood-based host response test for the prediction of bacterial and viral infections.

**Methods:** The PROMPT trial is a prospective, non-interventional, multi-center randomized, controlled clinical trial that enrolled 397 adult patients presenting to the ED with signs of acute infection and at least one vital sign change. The infection status was adjudicated using chart review (including a syndromic molecular respiratory panel, procalcitonin and C-reactive protein) by three infectious disease physicians blinded to InSep results. InSep (version BVN-2) was performed using PAXgene Blood RNA processed and quantified on NanoString nCounter SPRINT. InSep results (likelihood of bacterial and viral infection) were compared to the adjudicated infection status.

**Results:** Subject mean age was 64 years, comorbidities were significant for diabetes (17.1%), chronic obstructive pulmonary disease (13.6%), and severe neurological disease (6.8%); 16.9% of subjects were immunocompromised. Infections were adjudicated as bacterial (14.1%), viral (11.3%) and noninfected (0.25%): 74.1% of subjects were adjudicated as indeterminate. InSep distinguished bacterial vs. viral/noninfected patients and viral vs. bacterial/noninfected patients using consensus adjudication with AUROCs of 0.94 (95% CI 0.90-0.99) and 0.90 (95% CI 0.83-0.96), respectively. AUROCs for bacterial vs. viral/noninfected patients were 0.88 (95%CI 0.79-0.96) for PCT, 0.80 (95% CI 0.72-89) for CRP and 0.78 (95% CI 0.69-0.87) for white blood cell counts (of note, the latter biomarkers were provided as part of clinical adjudication). To enable clinical actionability, InSep incorporates score cutoffs to allocate patients into interpretation bands. The Very Likely (rule in) InSep bacterial band showed a specificity of 98% compared to 94% for the corresponding PCT band (>0.5 ug/L); the Very Unlikely (rule-out) band showed a sensitivity of 95% for InSep compared to 86% for PCT. For the detection of viral infections, InSep demonstrated a specificity of 93% for the Very Likely band (rule in) and a sensitivity of 96% for the Very Unlikely band (rule out).

**Conclusion:** InSep demonstrated high accuracy for predicting the presence of both bacterial and viral infections in ED patients with suspected acute infections or suspected sepsis. When translated into a rapid, point-of-care test, InSep will provide ED physicians with actionable results supporting early informed treatment decisions to improve patient outcomes while upholding antimicrobial stewardship.

**Take-home message:** InSep host response test is a point-of-care test providing with accuracy the likelihood for bacterial or viral infection for patients admitted at the emergencies InSep provided information on the likelihood of bacterial co-infection among patients with COVID-19.

## Introduction

The optimal approach to empiric antibiotic prescribing for suspected sepsis in the emergency department (ED), specifically timing and spectrum of therapy, is a matter of significant ongoing controversy[1-7]. Due to the fears of untreated bacterial illness and sepsis, clinicians are likely to default to a decision to prescribe antibiotics[1]. This pattern drives antibiotic overuse and resistance, despite considerable efforts to change behavior[2-4]. At present, the opportunity to tailor antibiotic therapy in EDs is limited by the lack of speed and sensitivity of culture-based techniques[5, 6].

The host response to infection can be measured on the cellular, protein and RNA level. White blood cell count, C-reactive protein and procalcitonin most often used to diagnose bacterial infections[5, 7] while there are no established markers for viral infections. Several European trials demonstrated that procalcitonin-guided management reduces antibiotic use[8, 9]; however, the incremental value of procalcitonin when used along best practice promotion of current guidelines has been questioned and a large US multicenter interventional trial failed to show a reduction in antibiotic use[10].

The InSep^™^ (Inflammatix, Bulingame, CA) acute infection and sepsis test is an innovative host-response assay which has the potential to address these unmet medical needs in acute infections and sepsis[11]. The assay measures 29 host mRNAs from peripheral blood and incorporates advanced machine learning to calculate three scores for predicting 1) the likelihood of bacterial infection, 2) the likelihood of viral infection, and 3) the risk for 30-day mortality[12-15]. Mayhew et al.[16] recently reported the first generation of a neural-network-based classifier for the 29-mRNA set (Inflammatix-Bacterial-Viral-Noninfected Version 1; IMX-BVN-1) applied in an independent cohort of patients enrolled within 36 hours of hospital admission; the classifier demonstrated high AUROCs of 0.92 (bacterial-vs.-other) and 0.91 (viral-vs.-other). The updated ‘BVN-2’ algorithm was detailed elsewhere[17].

The objective of the present study was to assess the accuracy of InSep for the detection of bacterial and viral infections using 3-physician clinical adjudication as the gold standard. We demonstrate that the InSep test accurately distinguishes bacterial and viral infections among ED patients in Greece (PROMPT study). In addition, we show the potential utility of InSep to detect bacterial co-infections in patients with confirmed SARS-CoV-2 infection enrolled using similar inclusion criteria at the same centers in 2020.

## Methods

### Patient enrollment

The PROMPT trial is a prospective, non-interventional, multi-center randomized, controlled clinical trial to assess the clinical validity of the heparin binding protein assay to indicate the presence and outcome of sepsis, including septic shock, in patients with suspected infection following emergency department admission. Patients were recruited from six sites in Greece participating in the Hellenic Sepsis Study Group (NCT 03295825, clinicaltrials.gov) between October 2017 and June 2018. The study was approved by the Ethics Committees of the participating hospitals. In a sub-group of this trial, PAXgene^®^ Blood RNA tubes were collected to assess the accuracy of the InSep test to distinguish between bacterial and viral infections. Enrolment was consecutive.

Screening for eligibility was done among patients admitted at the emergency departments of the participating hospitals. Inclusion criteria were: a) age ≥18 years; b) written informed consent provided by the patients or first-degree relatives; and c) suspicion of infection. Suspicion of infection was defined as the presence of at least one of the following: temperature > 38°C, temperature < 36°C, heart rate > 90 bpm, respiratory rate > 20/min, or self-reported fever/chills. No exclusion criteria applied. Full clinical information was recorded including demographics, severity scores, comorbidities, predisposing conditions and 28-day outcome. Blood was drawn after venipuncture of one forearm vein under aseptic conditions. One volume of 2.5 ml of blood was transferred into one PAXgene Blood RNA tube (PreAnalytics, Hombrechtikon, Switzerland) and kept refrigerated at −80^0^C; the remaining volume of 5 ml was transferred into one sterile and pyrogen-free tube and centrifuged.

In addition, between March and April 2020, 97 adults with confirmed molecular detection of SARS-CoV-2 in respiratory secretions and radiological evidence of lower respiratory tract involvement were enrolled at three study sites of the Hellenic Sepsis Study Group following approval of the Ethics Committees of the participating hospitals. PAXgene Blood RNA tubes were drawn within the first 24 hours from admission along with other standard laboratory parameters. Data collection included demographic information, clinical scores (SOFA, APACHE II), laboratory results, length of stay and clinical outcomes. Patients were followed up daily for 30 days to determine outcomes including respiratory failure (PaO2/FiO2 ratio less than 150 requiring mechanical ventilation) and/or death.

### Laboratory tests and result interpretation

Laboratory parameters including complete blood cell count and differential, biochemistry panel, blood gasses, microbiological (blood and urine culture) and viral tests were performed as indicated by local policies. C-reactive protein (CRP) and procalcitonin (PCT) were measured locally in serum by a nephelometric assay and a Kryptor assay, respectively.

A syndromic molecular respiratory PCR panels (FilmArray® Respiratory Panel, Biofire, Salt Lake City, UT) was performed for this study using frozen aliquots of nasopharyngeal swabs.

Positive results for microbiological and virological tests only considered clinically significant organisms, as determined by chart review and study-specific guidelines (Supplementary Methods). Negative results included patients with negative results, as well as those with positive results with no or uncertain clinical significance.

### The InSep acute infection and sepsis test

PAXgene Blood RNA samples were shipped to Inflammatix (Burlingame, CA, USA) and processed by technicians blinded to clinical outcomes. Briefly, RNA extraction from PAXgene Blood RNA was performed in batched mode using a standardized protocol on the QiaCube^®^ as previously described[16].

The InSep test consists of 29 target mRNAs composed of three separate, validated sub-panels: the 11-mRNA “Sepsis MetaScore”[13], 7-mRNA “Bacterial-Viral MetaScore”[14], and 11-mRNA “Stanford mortality score”[12], and has been described elsewhere[11]. RNA targets were counted using the NanoString nCounter^®^ SPRINT Profiler from 150 ng of isolated RNA; the expression of four housekeeping genes (*CDIPT, KPNA6, RREB1, YWHAB*) was also counted to normalize mRNA counts across samples[16]. In the current study, we applied the Bacterial Viral Non-infected (BVN)-2 algorithm of the InSep test[17]. The IMX-BVN-2 classifier was directly applied to the NanoString data, blinded to clinical outcomes.

### Clinical Adjudication

As there is no established gold standard to distinguish bacterial from viral infection clinical adjudication was performed to determine the ground truth, i.e. the presence of a bacterial and/or viral infection. Three independent, experienced, board-certified infectious disease physicians adjudicated all cases using a standardized medical chart derived from a case report form using an online electronic data capture system (Medrio). All cases were adjudicated into four pre-defined categories each for the presence of a bacterial and a viral infection as follows:

1. Yes: infection *proven* by clinical assessment plus results of microbiological or other laboratory tests.
2. Probable: Infection *likely* by clinical assessment but not confirmed by microbiological or other laboratory results.
3. Unlikely: Infection unlikely by clinical assessment but no other definite diagnosis established, or a microorganism detected but interpreted as colonization rather than cause of infection.
4. No: Infection ruled out by clinical assessment plus other definite diagnosis established

We calculated performance characteristics using two adjudication methods: consensus and forced. For consensus adjudication, only patients who were adjudicated as Yes/Yes/Yes, Yes/Yes/Probable, No/No/No or No/No/Unlikely by the three independent adjudicators for bacterial or viral infections (i.e. there was a “consensus” regarding infection status) were used in the performance evaluation. Patients with any other adjudication combination represent cases with an ‘indeterminate’ infection status and were removed from the consensus adjudication. However, in order to also evaluate the performance of InSep and comparator tests including all 397 patients we also determined the ‘forced adjudication’ where all cases leaning towards YES but not reaching consensus (e.g., Yes/Probably/Unlikely) were grouped together with those adjudicated as consensus Yes and cases leaning toward NO but lacking consensus were adjudicated as No.

Adjudicators were blinded to the InSep test results but had access to PCT and CRP results for their adjudication.

### Statistical analysis

Continuous variables are presented with the mean and standard deviation. Nominal variables are presented as n (% of group). The primary outcome of this study was the diagnostic performance of InSep bacterial and viral scores expressed as interpretation bands (Very Likely, Possible, Unlikely, Very Unlikely) when using ground truths (infection present or infection absent) established by the expert adjudicator panel. Performance metrics include the area under the receiver operating characteristic (AUROC), nominal likelihood ratios for each band, sensitivity and specificity for individual bands using routine statistical methods[18-20]. We also calculated the percentage of subjects that were allocated to each of the InSep interpretation bands.

## Results

We enrolled a total of 457 adult patients with suspected acute infection and at least one vital sign change across six EDs (Fig. 1); 30 patients denied participation, 27 were removed due to insufficient blood draws for the PAXgene Blood RNA tube, 2 were excluded due to missing clinical information and one patient was excluded due to errors during RNA extraction resulting in a final patient cohort of 397 patients.

**Fig 1.**
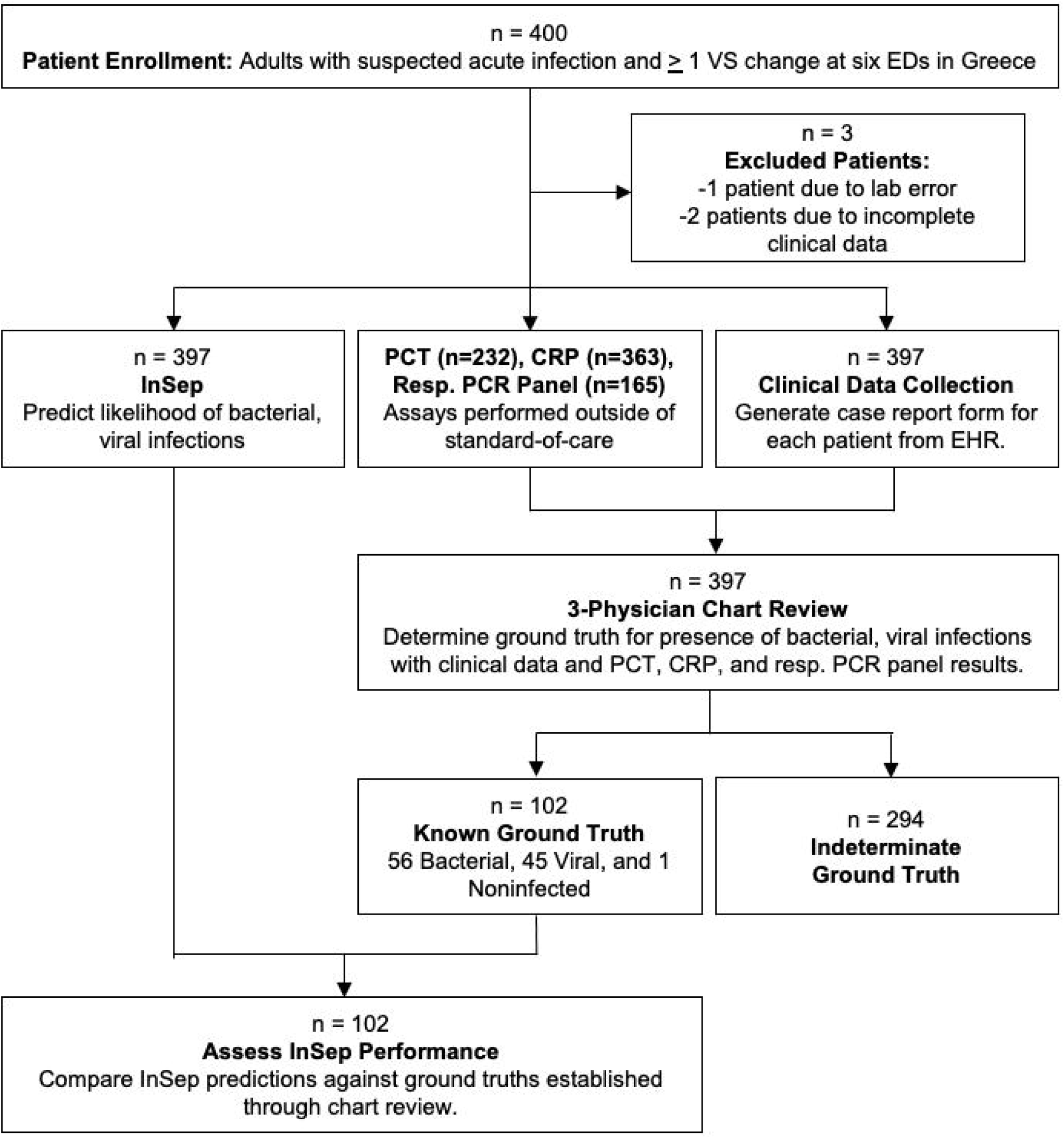
Study flowchart. 400 patients with suspected acute infection and ≥ 1 vital sign (VS) changes from six EDs in Greece were enrolled. Three physicians used electronic health records (EHRs) data alongside results from PCT, CRP, and respiratory syndromic panels to establish ground truths for the presence of bacterial and viral infections. InSep performance was was compared to ground truths.

Patient characteristics for the 397 patients enrolled are shown in Table 1. All patients were of Caucasian ethnicity. The most common comorbidities included type 1 or type 2 diabetes mellitus (68 patients, 17.1%), chronic obstructive pulmonary disease (54 patients, 13.6%), and severe neurological disease (27 patients, 6.8%). Sixty-seven patients (16.9%) were immunocompromised (45 with cancer or chemotherapy, 17 receiving steroids, 1 transplant recipient, 4 others). Two-hundred eighty-two patients (71.0%) were admitted to the hospital ward, 102 patients (25.7%) were discharged home, no patients were admitted to the ICU.

**Table 1.** Patient Characteristics.

| **Patient Characteristic** | **Number of Patients (%)**<br>**(n=397)** |
| --- | --- |
| Sex |  |
| Female | 203 (51.1%) |
| Male | 194 (48.9%) |
| Mean Age (Standard Deviation) | 62.1 (SD 21.9) |
| Ethnicity |  |
| White, Non-Hispanic | 397 (100%) |
| Immune Status |  |
| Immunocompetent | 330 (83.1%) |
| Immunocompromised | 63 (15.9%) |
| Unknown | 4 (0.010%) |
| Consensus Physician Adjudication |  |
| Bacterial | 56 (14.1%) |
| Viral | 45 (11.3%) |
| Noninfected | 1 (0.25%) |
| Indeterminant | 294 (74.1%) |
| Mortality within 30 days |  |
| Deceased | 38 (9.6%) |
| Survived | 359 (90.4%) |
| Subject Disposition |  |
| Discharged home | 102 (25.7%) |
| Ward | 282 (71.0%) |
| Telemetry/observation/step-down unit | 2 (0.50%) |
| Transferred out of hospital | 2 (0.50%) |
| Eloped | 1 (0.25%) |
| Unknown | 8 (2.0%) |
| Lab Parameters |  |
| WBC Mean (range, /mm<sup>3</sup>) | 11,385 (160-46,820) |
| CRP Mean (range, mg/l) | 66.2 (0.18-419) |
| PCT Mean (range, ng/ml) | 2.4 (0.02-126) |
| Comorbidities |  |
| Diabetes (Type I or Type II) | 68 (17.1%) |
| Chronic obstructive pulmonary disease | 54 (13.6%) |
| Congestive heart failure or pulmonary edema | 42 (10.6%) |
| Severe neurological disease | 27 (6.8%) |
| Chronic kidney disease with elevated creatinine | 24 (6.0%) |
| Rheumatoid arthritis | 6 (1.5%) |
| Major trauma or surgery within last 30 days | 5 (1.3%) |
| Other autoimmune diseases | 4 (1.0%) |
| Systemic Lupus erythematosus | 3 (0.76%) |
| Other comorbidities | 185 (46.6%) |
| None | 142 (35.8%) |
| Total immunocompromised patients | 67 (16.9%) |
| Steroids (due to comorbidities) | 20 (5.0%) |
| Cancer or chemotherapy | 45 (11.3%) |
| Solid organ transplant recipient | 1 (0.2%) |
| Other | 4 (1.0%) |

**Table 2.** Comparison of AUROCs applying consensus vs. forced adjudications for InSep and other commonly used biomarkers.

|  | **Consensus Adjudication**<br>**AUROC** | **Forced Adjudication**<br>**AUROC** |
| --- | --- | --- |
| **Bacterial vs. Viral/Noninfected** |  |  |
| InSep Bacterial Score | 0.94 (95% CI 0.90 - 0.99) | 0.76 (95% CI 0.71 - 0.81) |
| PCT | 0.88 (95% CI 0.79 - 0.96) | 0.77 (95% CI 0.70 - 0.83) |
| CRP | 0.80 (95% CI 0.72 - 0.89) | 0.69 (95% CI 0.63 - 0.74) |
| WBC | 0.78 (95% CI 0.69 - 0.87) | 0.70 (95% CI 0.64 - 0.75) |
| **Viral vs. Bacterial/Noninfected** |  |  |
| InSep Viral Score | 0.90 (95% CI 0.83 - 0.96) | 0.70 (95% CI 0.63 - 0.76) |

Physicians determined the ‘ground truth’ as follows: bacterial (56 patients, 14.1%), viral (45, 11.3%), noninfected (1, 0.25%), and indeterminate (294, 74.1%).

### InSep distinguishes bacterial from viral infections with high accuracy

InSep bacterial and viral scores showed an excellent ability to distinguish between patients with bacterial infections and those with viral infections or noninfectious etiologies (Fig. 2A) using consensus adjudication cases (ground truth); conversely, InSep was also able to accurately distinguish patients with viral infections from those with bacterial infection or non-infectious etiologies (Fig. 2B). InSep distinguished bacterial vs. viral/noninfected patients and viral vs. bacterial/noninfected patients with AUROCs of 0.94 (95% CI 0.90-0.99) and 0.90 (95% CI 0.83-0.96), respectively (Figure 2C-D). In comparison, AUROCs for bacterial vs. viral/noninfected patients for other biomarkers used to diagnose bacterial infections were as follows: PCT (0.88, 95%CI 0.79-0.96), CRP (0.80, 95% CI 0.72-89), and white blood cell counts (0.78, 95% CI 0.69-0.87) (Figure 2E).

**Fig 2.**
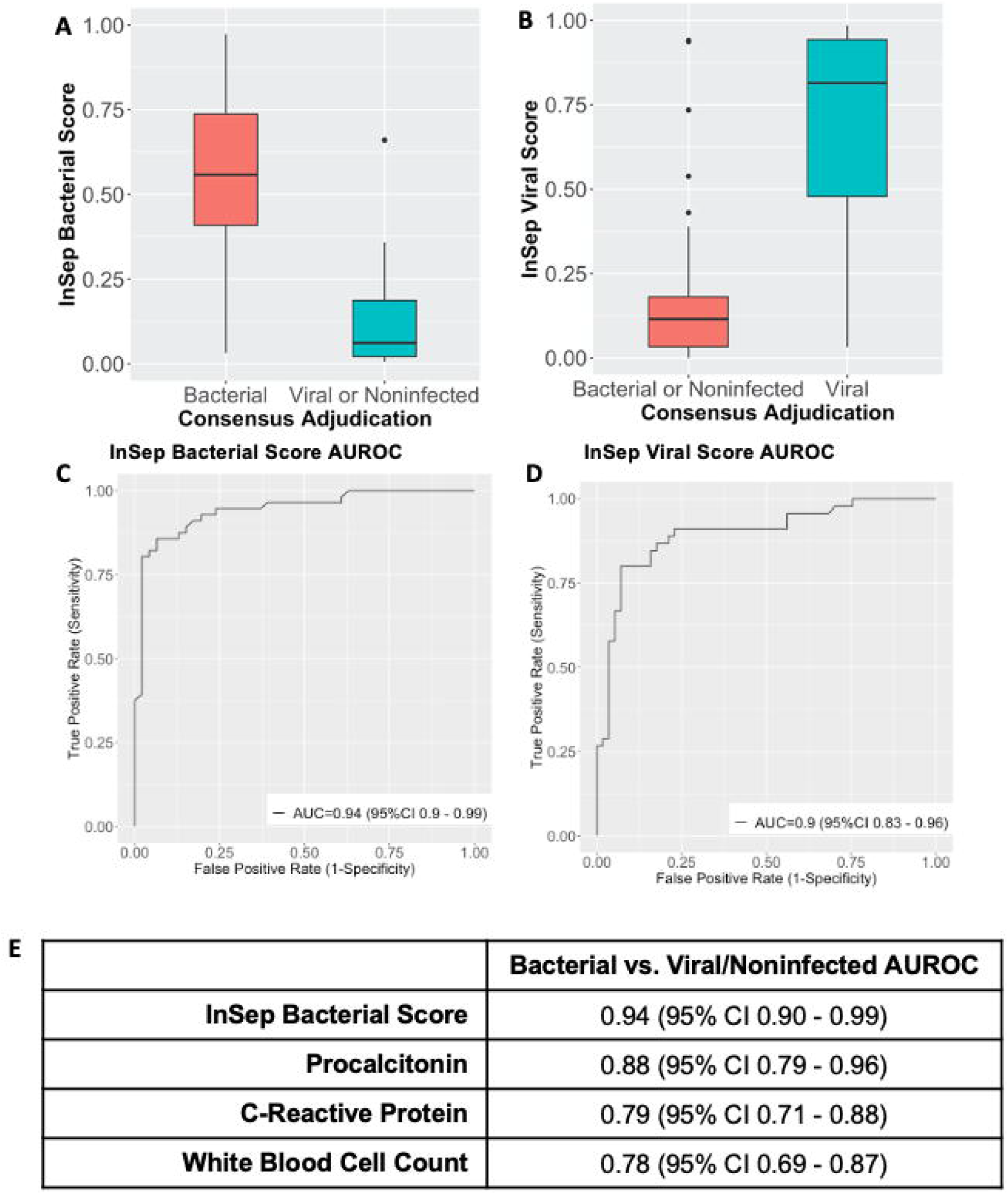
InSep testing accuracy using consensus adjudication. (A) InSep bacterial scores and (B) InSep viral scores among patients with known bacterial, viral, and non-infected status as determined by three-physician consensus adjudication. (C) Area under the receiver operating curves (AUROCs) for InSep bacterial scores (bacterial vs. viral/noninfected predictions). (D) Area under the receiver operating curves (AUROCs) for InSep viral scores (viral vs. bacterial/noninfected predictions). (E) Bacterial vs. viral/noninfected AUROCs for InSep bacterial scores, procalcitonin, C-reactive protein, and white blood cell counts.

### InSep performance using ‘forced adjudication’

While consensus adjudication is the “fairest” assessment for the accuracy of InSep (only including patients with highly confident infection status), it removes many patients due to ambiguity regarding infection status. Therefore, we also evaluated the performance of InSep using the forced adjudication which forces all Probable adjudications into a Yes-Probable category, and all Unlikely adjudications into a No-Unlikely category (see Methods section). Expectedly, AUROC’s for InSep, PCT, and CRP all decreased 14-19% upon applying the forced adjudication and ranged from 0.70 to 0.77 (Supplementary Table S1), reflecting the inaccuracies (lack of consensus) in the ‘forced adjudication’.

### InSep provides actionable results when segmented into interpretation (result) bands

While AUROCs are suitable for characterizing overall accuracy of diagnostic tests, they are not meaningful for individual patient management. Therefore, the InSep test provides absolute scores that fall into defined interpretation bands for the likelihood of a bacterial infection and the likelihood of a viral infection. Fig. 4A shows the results of InSep segmented into four interpretation bands (using previously established thresholds) for the likelihood of a bacterial infection compared to PCT (interpretation bands based on published data at concentrations of <0.1 ug/L, 0.1-0.25, >0.25-0.5 and >0.5[10], Figure 4B). The Very Likely (rule in) InSep band showed a specificity of 98% compared to 94% corresponding band for PCT (>0.5 ug/L). The Very Unlikely (rule-out) band showed a sensitivity of 95% for InSep compared to 86% for PCT. InSep results for the likelihood of a viral infection are presented in Fig. 4C. InSep demonstrated a specificity of 93% for the Very Likely band (rule in) and a sensitivity of 96% for the Very Unlikely band (rule out).

**Fig 3.**
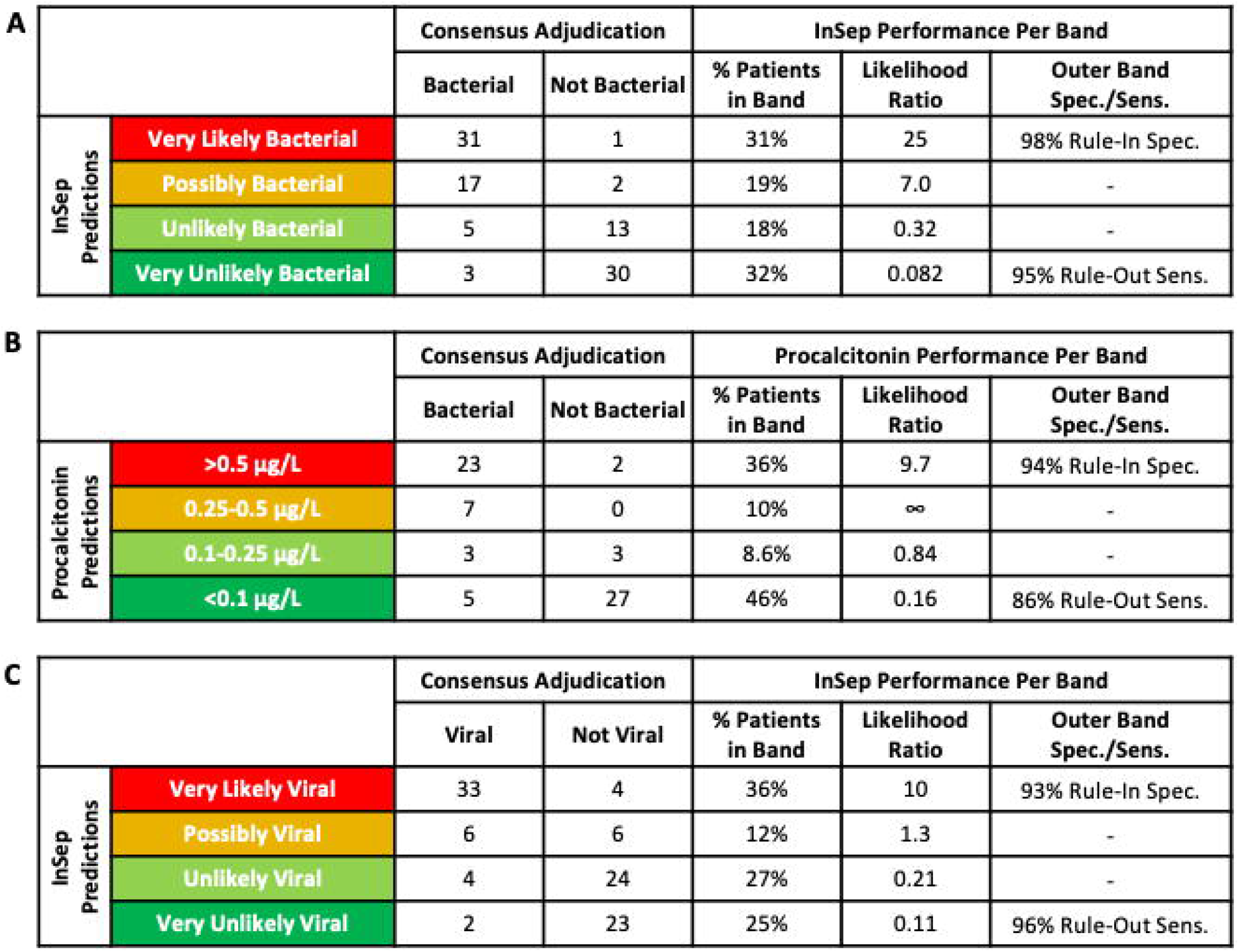
InSep interpretation bands provide clinically actionable results. (A) InSep bacterial results in interpretation bands ranging from Very likely, Possibly to Unlikely and Very unlikely. (B) Performance accuracy for procalcitonin depicted in interpretation bands for different concentration ranges reported in the literature[28]. (C) InSep viral results in interpretation bands ranging from Very likely, Possibly to Unlikely and Very unlikely.

**Fig 4.**
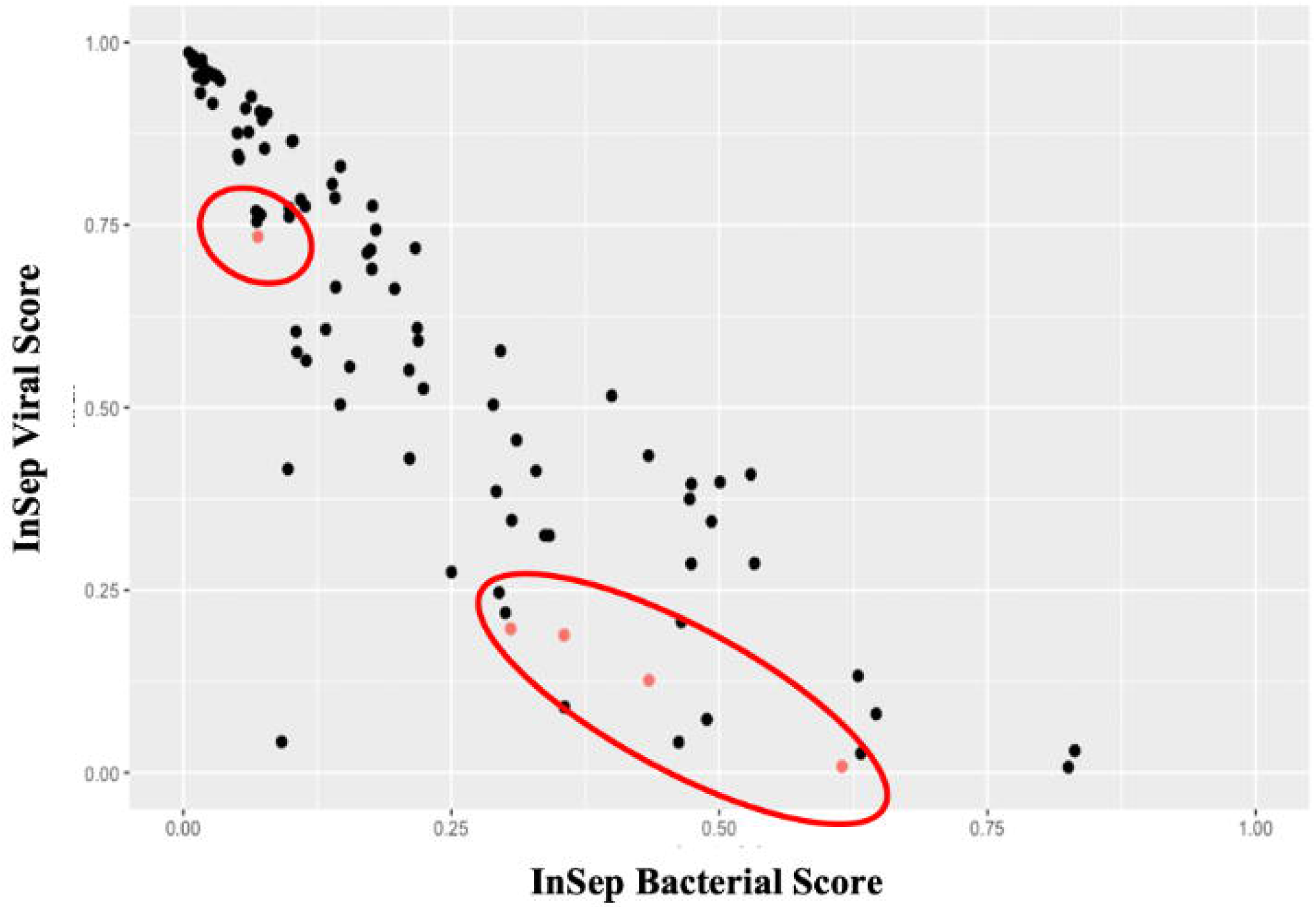
InSep bacterial (x-axis) and viral (y-axis) scores for 97 patients with confirmed SARS-CoV-2 infection. Patients with microbiologically confirmed bacterial (co-)infection are depicted in red (circled).

### InSep shows positive agreement with microbial test results

We also compared the performance of InSep against commonly used microbiological and virological tests to establish the presence of an infection (Supplementary Figure S1). Among 21 patients with positive and clinically significant blood culture results, InSep classified 15 patients as Very likely bacterial, 5 patients as Possibly bacterial, and 1 patient as Unlikely bacterial; there were no patients classified as Very unlikely bacterial. After excluding Possibly bacterial and Unlikely bacterial results, InSep showed 100% positive agreement compared to blood cultures. InSep showed a positive agreement of 88% compared to clinically relevant urine cultures, and a positive agreement of and 94% compared to results obtained for viral pathogens in respiratory syndromic panels (Supplementary Figure S1).

### InSep results do not appear to be affected by immune status of the patient

The InSep performance is expected to be consistent across patient sub-populations. Rather than testing for different AUROCs across many subgroups (which does not control for other variables) we instead assessed whether the InSep scores are significantly affected by patient characteristics. We developed two linear regression models using the InSep bacterial and viral scores as outcome variables and with age, sex, immunocompromise status, consensus adjudication, and lactate (used as a severity marker) as predictor variables (Supplementary Tables S1-S2). For both bacterial and viral scores, only adjudication status and lactate were significant predictors of InSep.

To explore InSep performance in immunocompromised patients we present descriptive statistics by showing InSep results segmented by interpretation bands and immune status in Supplementary Figure S3, without tests for significance due to small sample size (n=20 immunocompromised patients who have a consensus adjudication).

### InSep interpretation bands provide clinically actionable results

The IMX-BVN-2 algorithm incorporates established cutoffs to allow InSep bacterial and viral scores to be segmented into four interpretation bands ranging from Very likely to Possible, Unlikely and Very Unlikely (Figure 3). For both bacterial and viral results, more than 60% of patients had results in the informative Very unlikely and Very likely outer interpretation bands. Specificities of the Very likely interpretation bands for bacterial and viral InSep results were 98% and 93%, respectively, thereby providing actionable information for rule-in decisions. Similarly, sensitivities of the Very unlikely interpretation bands for bacterial and viral InSep results were 95% and 96%, respectively, thereby allowing for safe rule-out decisions.

Of the 33 patients that fell into the Very unlikely bacterial band, 3 were adjudicated as having bacterial infections. All three of these patients had positive urine cultures for *E. coli* consistent with acute pyelonephritis; two of these patients had negative blood cultures, no blood cultures were drawn in the third patient.

We observed one case in which the InSep result indicated a high likelihood of bacterial infection whereas the adjudication classified this patient as having a viral but not a bacterial infection. The patient was immunocompromised (cancer chemotherapy), had diabetes mellitus. He had very high CRP (120 mg/L) and procalcitonin concentrations (95 ng/mL). A syndromic respiratory viral panel revealed the presence of Coronavirus NL63 but no blood or urine cultures were taken. The patient was given IV antibiotics and was discharged after one day.

### InSep holds promise for detection of bacterial co-infection in subjects infected with SARS-CoV-2

Above, we demonstrated that InSep generates highly accurate results in ED patients with suspected infection. As the current SARS-CoV-2 pandemic is a major threat to EDs worldwide we also investigated the accuracy of InSep in a cohort of 97 patients diagnosed with SARS-CoV-2 admitted to the same EDs between March and April 2020.

As all patients were diagnosed with SARS-CoV-2 only one infection class (viral) is represented and AUROCs were therefore not calculated for InSep. However, five patients were found to have a microbiologically confirmed bacterial co-infection (urine antigen positive for *Streptococcus pneumoniae*, n=2; urine antigen positive for *Legionella pneumoniae*, n=1; syndromic respiratory panel positive for *Klebsiella pneumoniae* and *Moraxella catarrhalis*, n=1; sputum culture positive for *Staphylococcus aureus*, n=1). Of interest, all patients except for one (*S. aureus* in sputum culture) developed respiratory failure, and all five patients survived. The distribution of InSep scores is shown in Fig. 4. Four of five patients with bacterial co-infections had bacterial scores of >0.25 in the InSep test. The majority of patients had viral scores of >0.25; interestingly, many of the patients with low viral scores were found to have bacterial co-infections (red circle, Fig. 4). The one patient with a low bacterial score had a positive sputum culture for *S. aureus*; whether this is colonization or co-infection could not be adjudicated.

In comparison, PCT concentrations for 3 of the bacterial co-infections were < 0.1 ng/ml (the fourth patient had a PCT concentrations of 2.71 ng/ml; the 5^th^ subject with a confirmed co-infection did not have PCT measured). CRP concentrations ranged from 8.1 to 292.7 mg/l (82.9, 170.2, 216.0) in patients with bacterial co-infections and between 3.3 and 418 mg/l in the entire cohort. Thus, these bacterial co-infections could easily have been missed at the time of patient presentation in the ED using established biomarkers for the detection of bacterial infections.

## Discussion

Here we present the clinical validation of an innovative host immune mRNA expression test, InSep, which has the potential to improve the accuracy of diagnosis of acute infections and sepsis[11-13, 17, 21]. The cohort of 397 subjects enrolled in EDs in Greece, is representative of a European patient population with a mean age of 62, a high rate of comorbidities (>50%) and immunosuppression (17%). Subjects were adjudicated as bacterial (14.1%), viral (11.3%), or noninfected (0.25%) while the majority of subjects were considered indeterminant (74.1%). Using the consensus adjudication (excluding subjects considered indeterminant) InSep showed a very high AUROC of 0.94 for the detection of patients with acute bacterial infections vs. those with viral or no infection. Similarly, the AUROC for the detection of patients with viral infections vs. those with bacterial or no infections was 0.90. The observed accuracy of PCT, CRP and white blood cell counts (approved for the detection of bacterial infections) ranged from 0.78 to 0.88. The observation that InSep performs as well or outperforms other biomarkers is notable because the nature of our chart review introduced bias in favor of the other biomarkers: Physicians were provided with patients’ PCT, CRP and white blood cell count results during their chart reviews to establish patient infection status (bacterial, viral, non-infected). If PCT results that were not consistent with InSep influenced the physicians’ adjudications of a patient, then theoretically, infection status would more closely reflect PCT results than InSep results.

As there are no biomarkers for the detection of viral infections our finding of an AUROC of 0.90 is also of importance. Identification of patients with viral infections would allow early implementation of contact precautions measures and/or antiviral treatments (e.g., oseltamivir for influenza).

As expected, when using the forced adjudication (which included patients with indeterminate infection status in performance calculations) AUROC’s for InSep, PCT, and CRP all decreased markedly ranging from 0.70 to 0.77, reflecting the inaccuracies (lack of consensus) in the ‘forced adjudication’. While these ‘uncertain’ patients represent the most interesting group for improved diagnosis, it is very difficult to judge any biomarker’s performance in this group since the standard itself is in question.

Of importance, we observed that InSep results showed high agreement with results obtained in microbiological tests including blood culture. Positive agreement with blood culture findings was 100%, 94% with viral pathogen detection and 88% with clinically significant urine cultures. One immunocompromised patient had an InSep result indicating a high likelihood of bacterial infection although adjudicated to have a viral but not a bacterial infection; the possibility of bacterial co-infection could not be excluded.

Applying logistic regression, we found that for both InSep bacterial and viral scores, only the adjudication status and lactate but not age, sex, or immunosuppression were significant predictors of InSep results. Importantly, InSep results did not appear to be impacted by the immune status of the patient. As the proportion of immunocompromised patients in the present study is too low to draw statistical conclusion, the accuracy of InSep results will be assessed in larger studies and in studies specifically enrolling immunocompromised patients.

While the results presented above demonstrate the excellent accuracy of the InSep test, the management of individual patients in the ED requires test results that are clinically actionable. The InSep test results will be provided as a score for the likelihood of a bacterial infection, and separately as a score for the likelihood of a viral infection. Each score will fall into one of four interpretation bands ranging from very unlikely to unlikely, possible and very likely based on preset classifier cut-offs. This innovative result presentation allows the test to provide early rule-in (very likely) and rule out (very unlikely) interpretations addressing the unmet medical need to decide between early administration of antibiotics to prevent sepsis vs. antimicrobial stewardship[1, 22-24]. In support of safe rule-out decisions for bacterial and viral infections based on InSep results in the Very unlikely interpretation band, sensitivities were 95% and 96%, respectively; similarly, in support of rule-in decisions specificities of the Very likely interpretation bands for bacterial and viral InSep results were 98% and 93%, respectively. These results are strong indications that InSep will generate accurate and actionable results for rule-in and rule-out decisions. Of note, more than 60% of patients had (bacterial and viral likelihood) InSep results in the informative Very unlikely and Very likely outer interpretation bands thereby making InSep a test with high clinical value. Furthermore, results observed for InSep appear superior to those obtained with PCT in a direct comparison. While the clinical utility of PCT continues to be discussed controversely[10, 25] future trials comparing InSep to PCT will have to demonstrate the added value of InSep.

Lastly, InSep provides results for the likelihood of acute bacterial and also viral infection thereby allowing rapid detection of co-infections. While co-infections appear to be rare in subjects with COVID-19 the vast majority of these subjects still receive empiric antibiotic treatment making this an area of unmet medical need[26, 27]. We performed a preliminary investigation of the capability of InSep to detect bacterial co-infections in a cohort of subjects with SARS-CoV-2 infections at the same ED sites. We detected 1.5% co-infections using the InSep test at admission. This is slightly lower than the 3.5% reported in a recent meta-analysis[27]. However, the studies included in the meta-analysis used a variety of methods for the detection of bacterial infections (unspecified in many cases) and the percentage of co-infection ranged from 0 to 45%. InSep showed increased bacterial scores in four out of five COVID-19 patients with culture- or antigen-positive microbiology results from respiratory and/or urine tests. Interestingly, the one patient for which InSep did not show increased bacterial scores had *Staphylococcus aureus* detected from a sputum sample which could have been a colonization rather than an infection. Larger studies will have to confirm the value of InSep for the detection of bacterial co-infections to allow limiting empiric antibiotic therapy to patients with proven bacterial coinfection.

In the present study, the InSep test utilized the BVN-2 classifier for result generation. Details on the development of updated BVN classifiers have previously been discussed[16, 17]. InSep will be launched with the BVN-3 classifier based on data generated from recent independent patient cohorts. The test, run on the rapid Myrna™ instrument, will provide physicians with the likelihood of bacterial and viral infections as discussed here but will also provide a result for the severity of the condition (manuscript in preparation). We will also report on the implementation of the InSep test in ED departments when combining the utility as a diagnostic and prognostic test.

In conclusion, InSep demonstrated high accuracy for predicting the presence of both bacterial and viral infections in a large cohort of patients with suspected acute infections or suspected sepsis in the ED. The InSep test also showed promise for the detection of bacterial co-infections in COVID-19 patients. When translated into a rapid point-of-care test the InSep test holds promise to provide ED physicians with actionable results supporting informed treatment decisions early in the clinical course thereby improving patient outcomes while upholding antimicrobial stewardship.

## Data Availability

Data are available upon request to

## Acknowledgements

The authors would like to thank the patients who agreed to participate in the study. We also acknowledge the expert support by the staff at the collection sites, the clinical adjudicators Helen C. Stankiewicz Karita, MD, Miles Beaman, MD, and Silvan Vesenbeckh, MD, and the expert statistical support by James Wacker.

## Compliance with ethical standards

The PROMPT study was conducted under the approvals EBD2392/16.05.2017 of ATTIKON University General Hospital; 7/26.05.2017 of the General Hospital of Sparti; 1/20.07.2017 of the General Hospital of Syros; 8/12.07.2017 of the General Hospital of Halkida; 42550/20.10.2017 of the General Hospital of Argos; and 4/31.05.2017 of “G. Gennimatas” General Hospital of Thessaloniki

