## SUPPLEMENT for "A 29-mRNA Host Response Test from Blood Accurately Distinguishes Bacterial and Viral Infections Among Emergency Department Patients"

Affiliations:

ATTIKON University Hospital

1 Rimini Str

12462 Athens

Greece

**Supplementary Table S1** Linear Regression with InSep Bacterial Score as Dependent Variable.

Linear regression revealed that other than lactate the markers listed are not influencing the InSep score.

| Coefficients: | Estimate | Std. Error | t | P |  |
| --- | --- | --- | --- | --- | --- |
| (Intercept) | -0.007 | 0.102 | -0.069 | 0.945 |  |
| Consensus Adjudication: bacterial | 0.435 | 0.050 | 8.768 | 0.000 | *** |
| Age | 0.001 | 0.002 | 0.776 | 0.442 |  |
| Sex - Male | 0.021 | 0.045 | 0.459 | 0.648 |  |
| Immunocompromised - Yes | -0.066 | 0.047 | -1.391 | 0.171 |  |
| Lactate | 0.062 | 0.024 | 2.614 | 0.012 | * |

**Supplementary Table S2** Linear Regression with InSep Viral Score as Dependent Variable. Linear regression revealed that other than lactate the markers listed are not influencing the InSep score.

| Coefficients: | Estimate | Std. Error | t | P |  |
| --- | --- | --- | --- | --- | --- |
| (Intercept) | 0.227 | 0.154 | 1.473 | 0.148 |  |
| Consensus Adjudication: viral | 0.438 | 0.070 | 6.257 | 0.000 | *** |
| Age | 0.001 | 0.002 | 0.291 | 0.772 |  |
| Sex - Male | -0.022 | 0.062 | -0.355 | 0.724 |  |
| Immunocompromised - Yes | -0.012 | 0.065 | -0.188 | 0.852 |  |
| Lactate | -0.071 | 0.033 | -2.188 | 0.034 | * |

**Supplementary Figures**

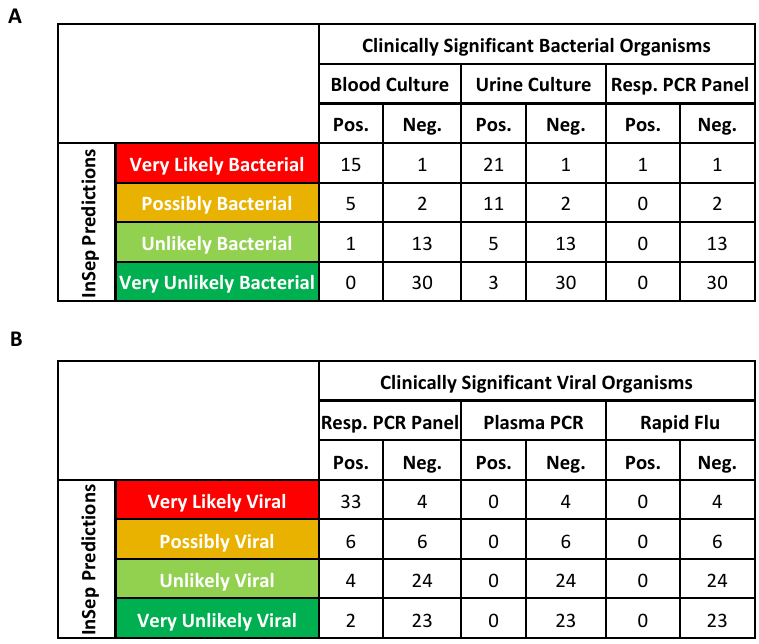

**Supplementary Fig. S1** InSep concordance with results of bacterial and viral pathogen detection tests. InSep results segmented by interpretation bands compared to conventional diagnostic test results for the presence of (A) bacterial and (B) viral pathogens. When considering only ‘Very likely’ results, InSep showed a positive agreement of 94% and 89% compared to positive bacterial cultures and viral findings in respiratory syndromic panels, respectively.

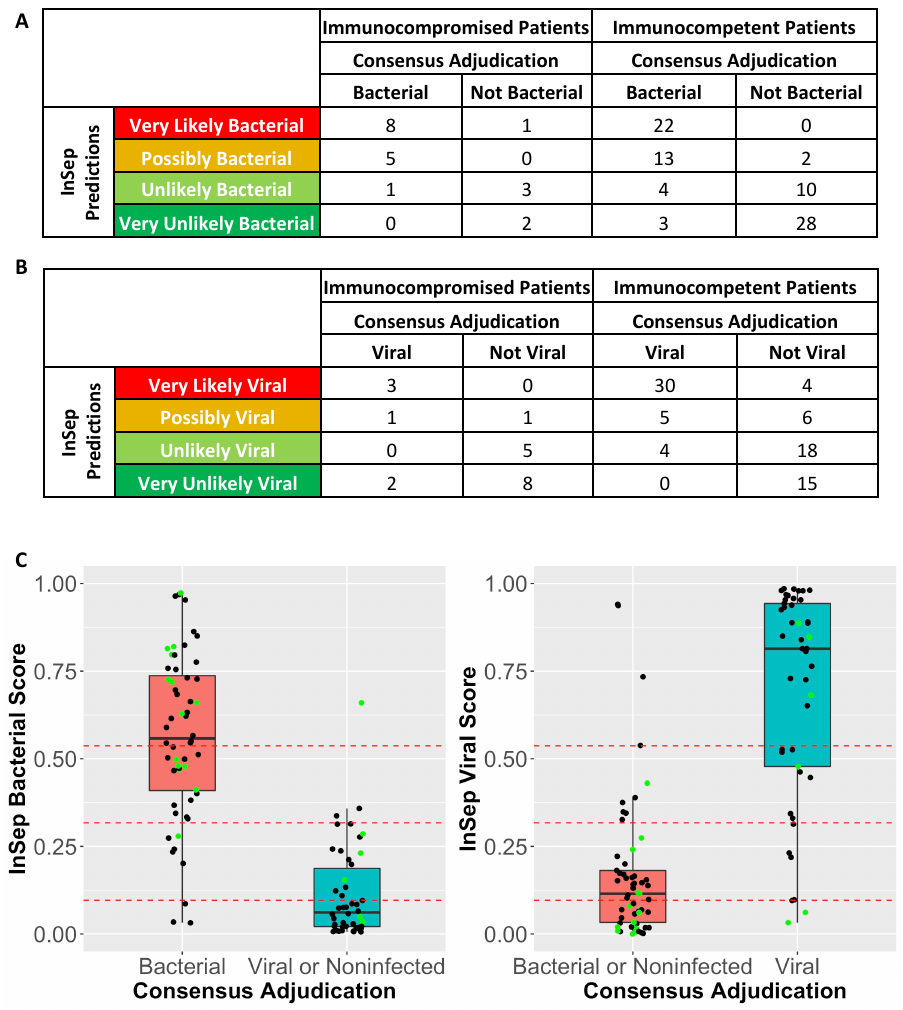

**Supplementary Fig. S2** InSep performance in immunocompromised patients. InSep (A) bacterial scores and (B) viral score results segmented by interpretation bands and immune status in the 20 (out of 102) consensus adjudicated patients who are immunocompromised. (C) shows the same boxplots as in Fig. 2) but with immunocompromised patients colored in green and dotted red lines which separate the four interpretation bands.
